# Deep Learning for Automated Detection of Generalized Paroxysmal Fast Activity in Lennox-Gastaut Syndrome

**DOI:** 10.1101/2023.05.21.23290211

**Authors:** Ewan S. Nurse, Linda J. Dalic, Shannon Clarke, Mark Cook, John Archer

## Abstract

**Objectives:** Generalized paroxysmal fast activity (GPFA) is a key EEG feature of Lennox-Gastaut Syndrome (LGS). Automated analysis of scalp EEG has been successful in detecting focal epileptiform abnormalities and generalized spike-wave discharges. Automatic detection of GPFA has been more challenging, due to its variability from patient to patient and similarity to normal brain rhythms such as sleep spindles. In this work, a deep learning convolutional neural network model is investigated for detection of GPFA events and estimating its overall burden from scalp EEG.

**Methods:** Data from 10 patients recorded during 4 ambulatory EEG monitoring sessions are used to generate and validate the model. All patients had confirmed LGS and were recruited into a trial for thalamic deep-brain stimulation therapy (ESTEL Trial).

**Results:** The correlation coefficient between manual and model estimates of event counts was r^2^=0.87 and for total burden (percentage of recording that is GPFA) was r^2^=0.91. Average GPFA detection sensitivity was 0.876, with an average false positive rate of 3.35 per minute. There was no significant difference found in the outcome measures between patients with early or delayed DBS treatment, an active VNS, indicating neuromodulation did not significantlly affect these results.

**Significance:** Overall, the deep learning model was able to accurately detect GPFA and provided accurate estimates of overall GPFA burden and electrographic event counts.

**Key points:**

- Deep learning has previously been shown to provide high-sensitivity seizure and inter-ictal automated event detection
- This work used a previously validated deep learning model to automatically detect GPFA in patients with LGS from scalp EEG
- Automated detections where highly correlated with human review for both number of events and burden of events

## Introduction

Lennox-Gastaut Syndrome (LGS) is a severe generalized epilepsy phenotype, characterized by multiple seizure types including tonic seizures, characteristic electroencephalographic (EEG) abnormalities (generalized paroxysmal fast activity (GPFA), slow-spike and wave (SSW)), and cognitive impairment^1,2^. A number of pharmacotherapies, vagal nerve stimulation (VNS), and corpus callosotomy^7^ are approved therapies for LGS, however patients rarely achieve seizure freedom. We, and others have investigated the use of deep-brain stimulation of the thalamic centromedian nucleus to reduce seizures in patients with LGS^8,9^.

GPFA may be an important electrographic feature of LGS, as it shows electrographic similarity to the initial phase of tonic seizures in LGS. GPFA occurs much more frequently in sleep, with a broad bilateral (‘generalized’) field consistent with its widespread cortical involvement ^2,4^. Our prior imaging^5^ and invasive studies^4^ suggest GPFA arises in distributed cortical networks, with propagation and amplification via deeper brain regions including brainstem, basal ganglia, and thalamus^6^.

Using visual inspection and manual mark-up, we have previously shown that changes in GPFA burden predict changes in the frequency of seizures recorded in seizure diaries in patients with LGS undergoing DBS ^3^. However, manual mark-up is laborious and potentially operator dependent. Hence, automated detection of GPFA to track the burden of these interictal discharges may be clinically useful for the ongoing management of LGS, especially as recording seizures in diaries can be difficult for certain patient groups^10,11^. Although seizure and interictal discharge detection software is becoming increasingly used in clinical practice ^12–15^, these systems have been optimized for common EEG abnormalities such as focal sharp waves and trains of generalized 3Hz spike and wave. The frequency band of GPFA is approximately 8-20 Hz, but varies from patient to patient. This overlaps with the frequency band of the posterior dominant rhythm and sleep spindles, creating challenges for frequency-based event detection.

GPFA occurs very frequently in LGS, averaging 1-5 discharges per minute^3^, meaning manual mark-up is highly time-consuming. A previous study generated a GPFA detector from EEG data recorded during EEG-fMRI in 13 children. This detector used time-frequency features with a variable threshold optimized to maximize the sensitivity (true-positive rate)^16^ and achieved a mean sensitivity of 61.3%, but with a very high false positivity rate.

In this work, we investigated the use of a deep convolutional neural network (CNN) model for estimating the event counts and burden of GPFA in ten patients with LGS. Adult patients with a prior diagnosis of LGS were enrolled into a study investigating DBS of the thalamic centromedian nucleus for treating seizures. Each patient undertook four 24h ambulatory EEG recordings at 12-week intervals, with the EEG manually marked for GPFA in a 2h window during sleep in each recording. We created a CNN to detect GPFA in the fourth EEG recording of each patient using a model architecture previously validated for measuring common EEG discharge types in idiopathic generalized epilepsy^14,15^. The CNN model is validated in two ways: comparing directly against the event count and burden, and more general detection metrics. Model performance is then compared between different DBS treatment groups, and whether patients had an active VNS.

## Methods

### Data Collection

The data used in this study was collected as part of our DBS therapy trial for LGS, described in full in previous publications^3,8^. Briefly, 20 adults with previously diagnosed LGS were recruited for a study of DBS to treat seizures. A subset of 10 of those cases with complete, annotated EEG records were used for this study. Participant identification numbers were not known to anyone outside the original study. Patients completed four ambulatory scalp EEG recordings at 12-week intervals. EEGs were collected with a standard 10-20 configuration, recorded at 256 Hz. This study uses recordings at four time points: at the time of implantation (week 0), and 12-, 24- and 36-weeks post implantation. At week 12, patients in the ‘early’ activation arm had their DBS turned on. At week 24 the remaining patients (‘delayed’ arm) had their devices activated. The week 0 and 12 data were used as model training, week 24 for validation, and 36 as held-out test data. This is visualized in Figure 1.

**Figure 1:**
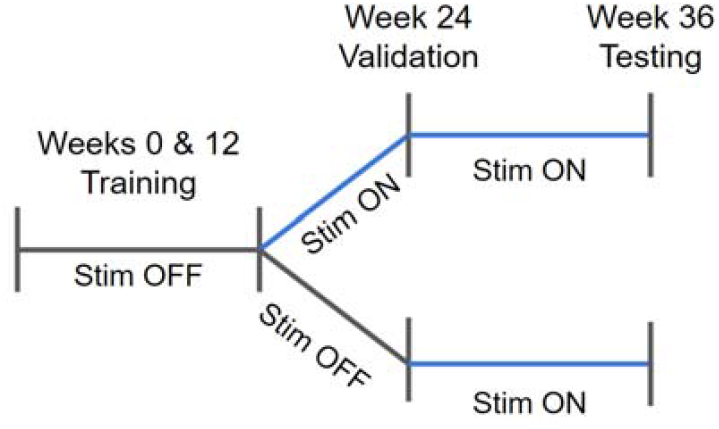
Study design. Participants had EEG recordings at 0, 12, 24, and 36 weeks post implantation. The ‘delayed’ group had their stimulation activated at 24 weeks, while the ‘early’ group were activated at 12 weeks. Data from weeks 0 and 12 were used for model training, week 24 for validation, and 36 for testing.

Table 1 shows patient demographics and GPFA counts. For each EEG, the trace was manually marked for GPFA between 00:00AM and 02:00AM. GPFA was defined as bursts of generalized fast epileptic activity of 10 – 30 Hz, with an amplitude greater than the background, lasting longer than 250 ms, but <5 s in duration.

**Table 1:** Patient demographics and event counts

| Pt | Sex (F/M) | Active VNS (Y/N) | DBS activation (Early/Delayed) | Training events – Wk 0 and 12 EEG (count) | Validation events – Wk 24 EEG (count) | Test events – Wk 36 EEG (count) |
| --- | --- | --- | --- | --- | --- | --- |
| E05 | F | Y | Delayed | 969 | 335 | 137 |
| E08 | F | N | Delayed | 1038 | 727 | 266 |
| E09 | F | N | Early | 1733 | 351 | 185 |
| E10 | F | N | Early | 1709 | 494 | 163 |
| E11 | F | Y | Early | 843 | 225 | 52 |
| E12 | M | N | Delayed | 1079 | 518 | 327 |
| E13 | M | N | Early | 3550 | 1444 | 343 |
| E14 | F | N | Early | 1252 | 292 | 278 |
| E18 | F | N | Delayed | 1031 | 488 | 415 |
| E19 | F | N | Early | 1349 | 749 | 814 |
| **Mean(std)** | - | - | - | 1455.3 (795.1) | 562.3 (355.1) | 298 (211.3) |
| **Total** | 80% F | 20% Y | 60% Early | 14553 | 5623 | 2980 |

**Table 2:** Secondary measures of model GPFA detection performance on testing data (weeks 24 and 36)

| Patient | Sensitivity (%) | False-positive rate (per minute) | Positive Predictive Value (%) | Data Reduction (%) |
| --- | --- | --- | --- | --- |
| E05 | 91.2 | 2.87 | 35.5 | 36.9 |
| E08 | 88.7 | 5.19 | 28.2 | 60.6 |
| E09 | 99.5 | 1.27 | 57.2 | 30.4 |
| E10 | 98.2 | 4.92 | 22.3 | 52.5 |
| E11 | 26.9 | 2.23 | 6.6 | 25.7 |
| E12 | 90.8 | 5.22 | 34.9 | 56.5 |
| E13 | 98.8 | 1.57 | 71.5 | 41.0 |
| E14 | 95.7 | 1.56 | 64.7 | 30.4 |
| E18 | 95.4 | 5.59 | 42.3 | 57.1 |
| E19 | 90.3 | 3.11 | 68.4 | 66.5 |
| **Average (std)** | **87.6 (20.5)** | **3.35 (1.6)** | **42.3 (22.65)** | **45.8 (13.8)** |

Two-sided t-tests were performed to assess the effect of stimulation devices (active VNS, or DBS early or delayed activation) on performance results (sensitivity, false-positive rate, PPV, and data reduction). No significant difference was found at $\alpha=0.05$ (with a Bonferroni correction).

### Detection Model

A convolutional neural network detection model was used that is structured similarly to the model used previously for IGE interictal discharge detection (14,16)). Briefly, the network consists of ‘blocks’ of convolutional, batch-norm and ReLU layers. The model consists of 20 blocks, described in Supplementary Table 1. The final layer is a single sigmoid that labels segments as GPFA if the output is over 0.5. All output labels less than 250 ms in duration were discarded. Any labels separated by less than 1s were merged into a single label. The network input has a receptive field of 427 samples at 256 Hz (1.67 seconds).

Network weights were initialized randomly. Stochastic optimization with a learning rate of 1e-3 and batch size of 80 was used to train the network. Each batch contained 50% positive samples (EEG segments labeled as GPFA) and 50% negative samples (any other segment of EEG). Networks were trained on NVIDIA Tesla M60 GPUs. EEG data were processed through three zero-phase Butterworth filters; a second order low-pass at 70 Hz, a second order high-pass at 0.5 Hz and a first order notch filter between 45 and 55 Hz (noting that line-noise in Australia occurs at 50Hz).

### Validation Measures

As the intended purpose of the model is to detect GPFA at a level equivalent to a human expert, the primary measures of model performance are the linear correlation between the expert and automated annotated total number of events and the total burden (percentage of EEG which is GPFA)^15^. Secondary measures are the sensitivity (percentage of GPFAs correctly detected), false-positive rate of detections, and data reduction (percentage of data that can be excluded from review assuming a 10 second EEG window during review). An example GPFA is shown in Figure 2.

**Figure 2:**
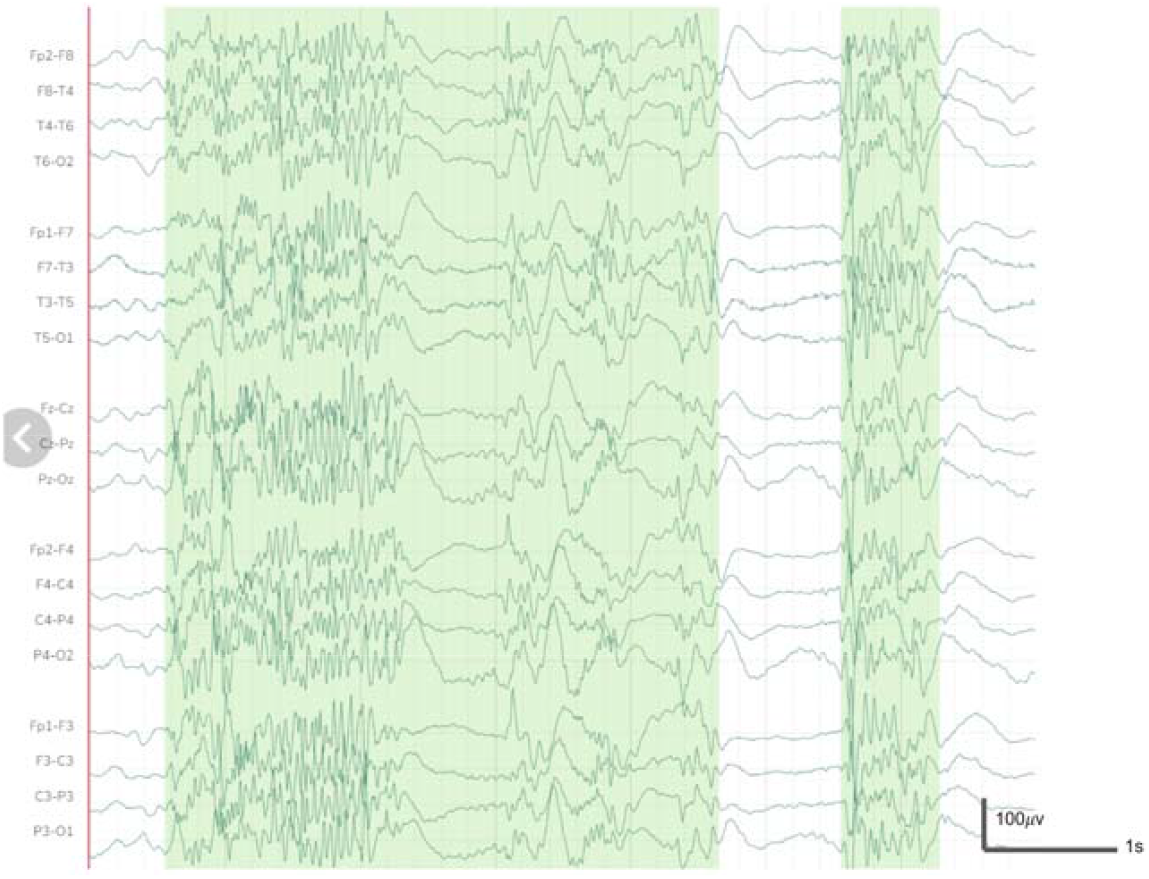
Typical example of a generalized paroxysmal fast activity events (green overlay) recorded by scalp EEG from sleep. Time base is 7 seconds, displayed with double-banana montage.

The effect of the VNS and DBS on classification performance was tested (two-sided t-test), as this has been previously shown to affect spectral features of scalp EEG in people with epilepsy, including those with LGS with leads implanted in the thalamic centromedian nucleus^17–19^.

## Results

### GPFA Event Count and Burden Estimation

Model performance for estimating the density and burden of GPFA compared to expert annotations are shown in Figure 3. Both number of events and burden are excellently correlated, with linear coefficients of determination (r^2^) values of 0.86 and 0.91, respectively. Therefore, the model is able to calculate GPFA burden in a given EEG study automatically with very similar accuracy to an expert reviewer.

**Figure 3.**
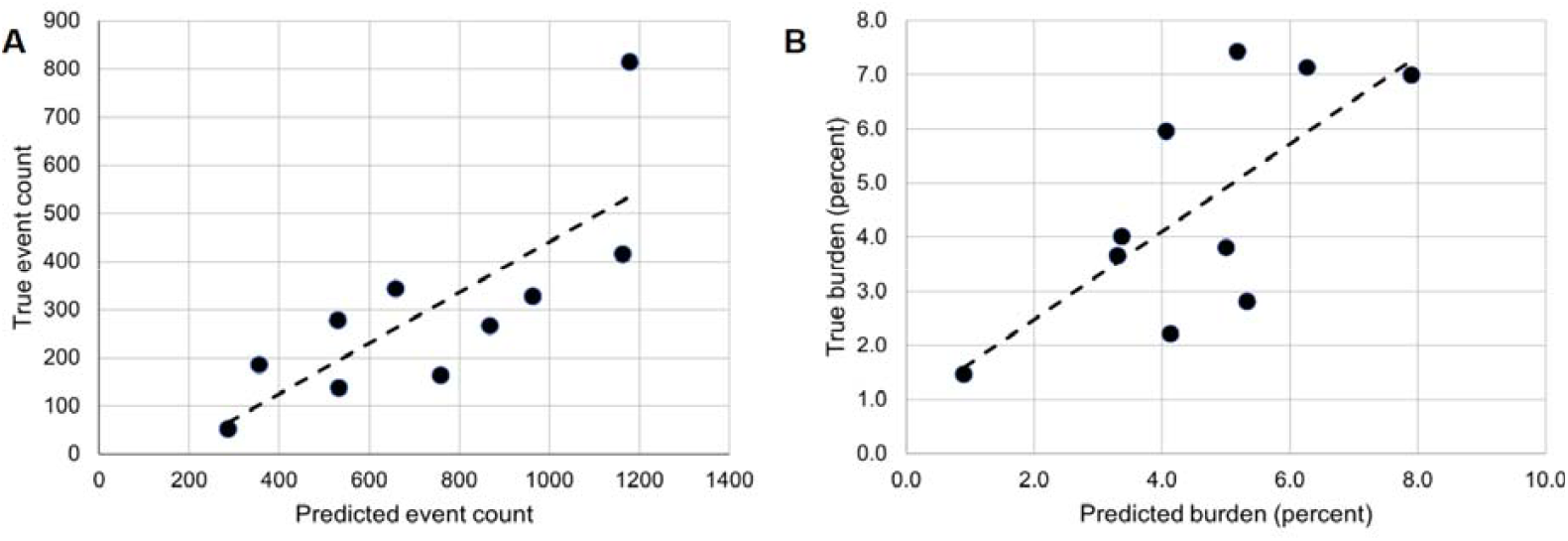
(a): Scatter plot of predicted against true count of labels per EEG study with linear line-of-best-fit (Dashed line) (p<0.0001, t-test) with excellent correlation (r^2^ = 0.87). (b): Scatter plot of predicted burden (total detected events duration divided by total recording duration) against true burden with linear line-of-best-fit (dashed line) (p<0.0001, t-test) with excellent correlation (r^2^ = 0.91).

### Model Performance

The average sensitivity and false-positive rate across the 10 test recordings are 0.876 and 3.35 events per minute, respectively. Full results are provided in Table 3. Most patients have a sensitivity of at least 0.9, indicating an excellent capability to detect true events, with the exception of E11. The false-positive rate of E11 was not an outlier, indicating that their GPFAs may be morphologically different to the rest of the cohort. The mean data reduction after labeling was 45.8%, indicating that review assisted by this model would on average nearly reduce review time by a half, while only missing <15% of true GPFA events.

## Discussion

Automation of EEG review is a topic of increasing investigation^12,13^, particularly for continuous recordings of weeks to months in duration^20^. Long-term automated counting of GPFA events may have potential benefit for the prediction of seizure events or titration of therapies, as has been demonstrated in other epilepsies^26–28^. Furthermore, there is increasing interest in the use of EEG measurements as trial endpoints for studying the effects of interventions, which would benefit immensely from automated review processes^21^.

The GPFA detection model presented in this work demonstrated high correlation with an expert reviewer in terms of both absolute event count and total discharge burden. Furthermore, the average sensitivity was close to 90%, indicating that the model could identify positive EEG segments with high accuracy. Interestingly, this is a similar level of performance to how this model architecture previously performed on IGE discharge detection, with an average sensitivity of about 90%^14,15^.

No performance decline was noted between patients in the ‘early’ or ‘delayed’ stimulation groups. This could be due to the changes in the cortical EEG being imperceivable to the model, or that the differences were suitably learned from the validation data that contained some recordings with active stimulation. Further analysis of cortical EEG before and after DBS activation could be undertaken to better characterize any potential changes.

A previous model created specifically for GPFA detection^16^ achieved a sensitivity of 61.3%, substantially less than in this work. This is likely due to a combination of factors: the use of a simple model with prescriptive feature engineering, and training on substantially less data per patient. For a number of detection problems, it is clear that deep-learning frameworks are superior to hand-coded features, and this seems to hold true for EEG abnormality detection^22,23^. Deep learning typically requires a large corpus of data though, and may not be ideal for creating models from smaller research datasets^24^.

The model created for this work exclusively sought to distinguish GPFA from any other EEG segment. The concept of ‘false-positive’ may be poorly defined, as a non-GPFA segment may still contain EEG that is otherwise abnormal, such as slow-spike wave^25^. Future work will seek to use multi-class models that can distinguish between different types of EEG events.

The model used in this study could be potentially improved in a number of ways. Patient-specific modeling may be more computationally expensive, but give better overall results, as individual morphologies may subtly differ. The model architecture used is also naive to the temporal nature of EEG, and models such as long short-term memory (LSTM) networks may provide higher performance by accounting for this^29,30^. No feature engineering was undertaken in this work, which may potentially be of benefit due to the well specified definition of GPFA. A downside to the deep learning approach is the high volume of data required to train the model compared to simpler feature engineering approaches, as well as the relatively limited capacity to learn about features of the EEG from the relatively ‘black box’ model structure.

A limitation of this work is that the GPFA annotations were provided by only a single expert reviewer. Having annotations from multiple markers would increase the generalizability of the proposed methods.

## Conclusion

This study has demonstrated that GPFA can be automatically detected with high sensitivity from scalp EEG using a deep learning approach. Automated detections had a high correlation to the event count and total GPFA burden. GPFA detection may be useful for future research, as well as automated diagnosis or treatment systems.

## Data Availability

All data produced in the present study are available upon reasonable request to the authors

## Funding Acknowledgement

This project was supported by NHMRC project grant 1108881

## Ethical Statement

This study received institutional approval from Austin Health Human Research Ethics Committee prior to trial commencement (approval number HREC/16/Austin/139).

We confirm that we have read the Journal’s position on issues involved in ethical publication and affirm that this report is consistent with those guidelines.

## Authorship Statement

ESN: Conceptualization, methodology, analysis, software, manuscript preparation

LJD: Conceptualization, data curation, analysis, manuscript preparation

SC: Methodology, software, analysis, manuscript preparation

MC: Analysis, supervision, manuscript preparation

JA: Conceptualization, supervision, manuscript preparation

