## Supplemental Table S1 for "Deep Learning for Automated Detection of Generalized Paroxysmal Fast Activity in Lennox-Gastaut Syndrome"

### Supplementary

**Table S1:** CNN model structure

| Block Number | Convolution Size |
| --- | --- |
| 1 | 64 |
| 2 | 64 |
| 3 | 64 |
| 4 | 64 |
| 5 | 128 |
| 6 | 128 |
| 7 | 128 |
| 8 | 256 |
| 9 | 256 |
| 10 | 256 |
| 11 | 512 |
| 12 | 512 |
| 13 | 512 |
| 14 | 256 |
| 15 | 128 |
| 16 | 128 |
| 17 | 128 |
| 18 | 128 |
| 19 | 128 |
| 20 | 128 |
